# Multi-Cancer Early Detection Tests: National Estimates of Awareness and Perceived Value in the United States

**DOI:** 10.1101/2025.08.02.25332855

**Authors:** Jemar R. Bather, Melody S. Goodman, Daniel Chavez-Yenter, José A. Pagán, Kimberly A. Kaphingst

**Affiliations:** Center for Anti-racism, Social Justice & Public Health, New York University School of Global Public Health, New York, NY, USA; Department of Biostatistics, New York University School of Global Public Health, New York, NY, USA; Division of Hematology Oncology, Perelman School of Medicine at the University of Pennsylvania, Philadelphia, PA, USA; Department of Medical Ethics and Health Policy, Perelman School of Medicine at the University of Pennsylvania, Philadelphia, PA, USA; Department of Public Health Policy and Management, New York University School of Global Public Health, New York, NY, USA; Huntsman Cancer Institute, Salt Lake City, UT, USA; Department of Communication, University of Utah, Salt Lake City, UT, USA

**Keywords:** medical screening, cancer prevention, cancer control, medical testing, liquid biopsy, diagnostic test, mass screening

## Abstract

**Background:** Multi-cancer early detection (MCED) tests represent a promising advancement in cancer screening technology. These tests can identify biomarkers from multiple cancer types using a single biological sample, potentially reducing cancer mortality and addressing cancer inequities. Little is known about MCED test awareness and perceived value among adults in the United States (U.S.).

**Methods:** Using nationally representative data from the 2024 Health Information National Trends Survey, we computed national estimates of MCED test awareness and perceived value and evaluated whether estimates varied across different population groups.

**Results:** The weighted sample represented 244 million U.S. adults, with 16.8% (95% CI: 15.2%-18.3%) aware of MCED tests and 42.1% (40.0%-44.2%) perceiving them as very valuable. In adjusted analyses, Hispanic ethnicity (adjusted prevalence ratio [aPR] = 1.29 [1.01-1.65]) and health-related social media use (aPR = 1.04 [1.01-1.06]) were associated with MCED test awareness. Factors associated with perceiving MCED tests as very valuable included: older age (e.g., 35-49 vs. 18-34: aPR = 1.43 [1.20-1.70]), minoritized race/ethnicity (non-Hispanic Black: aPR = 1.31 [1.15-1.50]; Hispanic: aPR = 1.40 [1.22-1.60]), family cancer history (aPR = 1.17 [1.03-1.34]), and frequent patient portal use (≥6 times vs. none: aPR = 1.23 [1.06-1.42]).

**Conclusion:** While awareness remains low, high perceived value among older adults and minoritized racial/ethnic populations suggests readiness for MCED adoption in groups that may benefit most from early cancer detection.

**Impact:** This report provides baseline data to inform equitable access strategies as MCED tests await approval from the U.S. Food and Drug Administration and seek coverage from insurers.

**Condensed abstract:** Only 16.8% of U.S. adults were aware of multi-cancer early detection tests in 2024, yet 42.1% perceived them as very valuable, with higher value perception among older adults, minoritized racial/ethnic populations, and those with family cancer history. These findings highlight both a substantial awareness-value gap and strong potential demand among groups that may benefit most from early cancer detection, providing critical baseline data to inform equitable implementation strategies as MCED tests await FDA approval and insurance coverage.

## Introduction

Cancer remains a leading cause of death in the United States (U.S.), with over 600,000 deaths projected for 2025 (1). Nearly 60% of these deaths will result from cancers lacking established screening protocols, including ovarian and liver cancers (1). Multi-cancer early detection (MCED) tests are a novel technology that represent a promising advancement in cancer screening for which approval is pending by the U.S. Food and Drug Administration (FDA) (2–5). These tests can identify biomarkers from multiple cancer types using a single biological sample (2–5), potentially reducing cancer mortality and addressing cancer inequities between privileged and disadvantaged populations (3,6). However, despite their potential impact, little is known about MCED test awareness and perceived value among U.S. adults. Understanding these factors is critical for the equitable implementation of this emerging technology (2,3).

Prior research suggests high interest in MCED testing among specific populations (7–9). A study in a Philadelphia health system found that 79% of primary care patients aged 50-80 years reported high interest in MCED testing (8). Similarly, 72% of U.S. adults aged 50-80 years preferred receiving MCED tests in addition to standard cancer screenings (9), and 75% of U.S. adults aged 45-80 years would undergo testing if free or covered by insurance (7). While these studies suggest strong potential demand for MCED testing (7–9), they have limitations. All of them excluded younger adults (7–9), a critical gap given recent shifts toward earlier screening ages for colorectal and breast cancers (10,11). Additionally, these studies focused on hypothetical interest rather than actual awareness or perceived value. This leaves a critical knowledge gap, as no nationally representative data sources exist on MCED test awareness across all adult age groups.

Currently, MCED tests cost approximately $949 and are not covered by most health insurance plans (2,3), creating a scenario where only affluent populations can access this potentially life-saving technology. Without health insurance coverage, this promising cancer screening tool risks widening the cancer inequities it could help eliminate (3,5,6). Therefore, establishing national estimates of MCED test awareness and perceived value is essential to inform health insurance coverage decisions, guide stakeholder engagement, and develop targeted public health campaigns.

Using data from the 2024 Health Information National Trends Survey (HINTS), we computed national estimates of MCED test awareness and perceived value and evaluated whether these estimates varied significantly across different population groups. We hypothesized that awareness would be low (<20%) given that MCED testing is an emerging health technology (2,3). We also hypothesized that MCED test awareness would be significantly associated with older age, non-Hispanic White identification, higher socioeconomic status (e.g., college graduate, high income), having a personal or family cancer history, living in an urban area, and greater patient portal use based on patterns observed in genetic testing research (12–21). For perceived value, we hypothesized that higher value perception would be observed among individuals with a family cancer history compared to those without a family cancer history.

## Materials and Methods

### Study Design and Setting

This cross-sectional study analyzed data from the HINTS 7, a nationally representative survey conducted by the National Cancer Institute from March 25 to September 16, 2024 (22–24). HINTS 7 assessed how U.S. adults access and use cancer-related information across the cancer control continuum, including prevention, diagnosis, and survivorship. The survey sampled noninstitutionalized U.S. adults aged 18 years or older using a probability-based design.

The Marketing Systems Group provided a sampling frame of U.S. addresses. Within each sampled household, one adult was selected using the next-birthday method, whereby the adult with the next upcoming birthday was asked to complete the questionnaire. Data were collected using a multi-mode approach following a modified Dillman protocol (25). Respondents could complete either a paper questionnaire or web-based survey. The initial mailing included a paper questionnaire with instructions for the web option, followed by a reminder postcard and two follow-up mailings. Households identified as potentially Spanish-speaking received materials and questionnaires in both Spanish and English.

The weighted response rate was 27.3% according to the Association of Public Opinion Research’s Response Rate 4 formula (26). Survey weights accounted for nonresponse bias and coverage error using calibration adjustments derived from the American Community Survey (age, sex, education, race/ethnicity, and Census region) and the National Health Interview Survey (health insurance coverage and cancer status). Additional information about the HINTS 7 study protocol is available elsewhere (26). Of 7,278 completed surveys, we restricted the unweighted analytic sample to 6,709 (92.2%) respondents with complete outcome data. This secondary analysis of publicly available data did not require review from the New York University Institutional Review Board. We followed the Strengthening the Reporting of Observational Studies in Epidemiology guidelines (27).

### Dependent Variables

HINTS 7 introduced two items assessing MCED test awareness and perceived value (26). Respondents were shown the following text: “Scientists have developed new tests to ‘screen’ for cancers early when they are easier to treat. These new tests, called Multi-Cancer Early Detection tests, use a single blood test to detect many different cancers at the same time.” The first item asked participants, “Before today had you ever heard of Multi-Cancer Early Detection tests?” We analyzed this as a binary outcome (1 = Yes, 0 = No). Participants rated their personal value of MCED testing with the question: “How valuable do you think it would be for you to have a Multi-Cancer Early Detection test right now?” Response options ranged from *not at all valuable* to *very valuable* on a 4-point scale. We dichotomized responses as very valuable versus all other categories.

### Independent Variables

#### Demographic & Socioeconomic Characteristics

We analyzed self-reported demographic and socioeconomic variables. Age was categorized into five groups: 18-34, 35-49, 50-64, 65-74, or ≥75 years. Sex assigned at birth was measured as male or female. Race/ethnicity included non-Hispanic White, non-Hispanic Black, Hispanic, non-Hispanic Asian, or non-Hispanic Other (including American Indian/Alaska Native, Native Hawaiian/Pacific Islander, and multiracial respondents). Educational attainment was classified as less than high school, high school graduate, some college, or college graduate or higher. Annual household income was categorized as <$50,000, $50,000-$99,999, or ≥$100,000. Additional socioeconomic indicators included health insurance coverage (yes, no), geographic urbanicity (urban, rural), and census region (Northeast, Midwest, South, West). Urbanicity was determined using the U.S. Department of Agriculture’s 2010 Rural-Urban Commuting Area codes (28).

#### Health Status & Medical History

We assessed personal and family cancer history through self-report. Respondents indicated whether they had ever been diagnosed with cancer (yes, no) and whether any first- or second-degree biological relative had cancer (yes, no, not sure). Chronic health conditions were measured by asking: “Has a doctor or other health professional ever told you that you had any of the following medical conditions?” The conditions included: (a) diabetes or high blood sugar; (b) high blood pressure or hypertension; (c) a heart condition such as heart attack, angina, or congestive heart failure; (d) chronic lung disease, asthma, emphysema, or chronic bronchitis; and (e) depression or anxiety disorder. We summed affirmative responses to create a chronic condition count (0, 1, or ≥2).

#### Health Information Behavior & Technology Use

Patient portal use was assessed with the question: “How many times did you access your online medical record or patient portal in the last 12 months?” Response options (0, 1-2 times, 3-5 times, 6-9 times, or ≥10 times) were consolidated into three categories: 0, 1-5 times, or ≥6 times. Health-related social media use was measured using five items with the following prompt: “Sometimes people use the internet to connect with other people online through social media. Examples of social media sites include Facebook, TikTok, YouTube, and Instagram. In the past 12 months, how often did you do the following?” (a) visited a social media site; (b) shared personal health information on social media; (c) shared general health-related information on social media (for example, a news article); (d) interacted with people who have similar health or medical issues on social media or online forums; and (e) watched a health-related video on a social media site (for example, YouTube). Responses were recorded on a 5-point scale from “never” to “almost every day”. Items were summed to create a composite score (Cronbach’s α = 0.71), with higher scores indicating greater health-related social media use (29).

### Statistical Analysis

All analyses were conducted using R version 4.5.1 (R Core Team, R Foundation for Statistical Computing). We accounted for HINTS 7’s complex survey design using the survey R package (30) and generated weighted summary statistics with the gtsummary R package (31). We examined bivariate relationships of each independent variable with MCED awareness and perceived value using design-adjusted Pearson’s chi-squared tests for categorical variables and Kruskal-Wallis tests for continuous measures. For multivariable analyses (32), we used modified Poisson regression with generalized estimating equations to estimate adjusted prevalence ratios (aPRs) and 95% confidence intervals (CIs) for factors associated with each outcome (33,34). To address missing data, we employed multiple imputation by chained equations using the mice R package (35). Imputation models included predictive mean matching for continuous variables, logistic regression for binary variables, multinomial logistic regression for nominal variables, and ordinal logistic regression for ordered categorical variables. We generated ten imputed datasets and pooled results using Rubin’s rules (36). Statistical significance was set at *p*<0.05.

## Results

### Weighted Sample Characteristics and Bivariate Analyses

The weighted sample represented 244 million U.S. adults (Table 1). The population was evenly distributed by age (50.9% under 50 years) and sex (50.4% male), with 58.6% identifying as non-Hispanic White (Table 1). Overall, 16.8% (95% CI: 15.2%-18.3%) of U.S. adults were aware of MCED tests, and 42.1% (95% CI: 40.0%-44.2%) perceived MCED testing as very valuable. MCED awareness varied significantly by education and health-related social media use. Adults with college degrees comprised 36.9% of the aware group, compared to 33.0% of the unaware group (*p*=0.042). Those aware of MCED tests (mean = 11.0) more frequently used social media for health-related purposes than individuals who were unaware of MCED tests (mean = 10.4, *p*<0.001).

**Table 1.** Weighted summary statistics, overall and by awareness of multi-cancer early detection tests in the United States, 2024 Health Information National Trends Survey.

| Characteristic | Aware of Multi-Cancer Early Detection tests |  |  | p-value |
| --- | --- | --- | --- | --- |
|  | Overall<br>N = 243,999,920 | No<br>N = 203,070,440 (83.2%) | Yes<br>N = 40,929,480 (16.8%) |  |
| **Perceived Multi-Cancer Early Detection testing as very valuable, No. (%)** |  |  |  | **0.90** |
| No | 141,250,363 (57.9) | 117,656,832 (57.9) | 23,593,531 (57.6) |  |
| Yes | 102,749,557 (42.1) | 85,413,608 (42.1) | 17,335,949 (42.4) |  |
| **Age, No. (%)** |  |  |  | **0.50** |
| 18-34 | 60,987,557 (25.0) | 50,439,157 (24.8) | 10,548,399 (25.8) |  |
| 35-49 | 63,198,434 (25.9) | 53,995,660 (26.6) | 9,202,773 (22.5) |  |
| 50-64 | 63,566,758 (26.1) | 52,031,310 (25.6) | 11,535,448 (28.2) |  |
| 65-74 | 31,483,424 (12.9) | 26,177,900 (12.9) | 5,305,524 (13.0) |  |
| 75+ | 21,567,415 (8.8) | 18,071,796 (8.9) | 3,495,618 (8.5) |  |
| Missing | 3,196,333 (1.3) | 2,354,615 (1.2) | 841,718 (2.1) |  |
| **Sex assigned at birth, No. (%)** |  |  |  | **0.20** |
| Male | 122,953,234 (50.4) | 101,603,127 (50.0) | 21,350,106 (52.2) |  |
| Female | 117,326,324 (48.1) | 98,831,308 (48.7) | 18,495,016 (45.2) |  |
| Missing | 3,720,363 (1.5) | 2,636,005 (1.3) | 1,084,358 (2.6) |  |
| **Race/ethnicity, No. (%)** |  |  |  | **0.40** |
| non-Hispanic White | 142,865,301 (58.6) | 119,644,845 (58.9) | 23,220,456 (56.7) |  |
| non-Hispanic Black | 25,961,569 (10.6) | 21,424,744 (10.6) | 4,536,825 (11.1) |  |
| Hispanic | 41,284,739 (16.9) | 33,055,471 (16.3) | 8,229,268 (20.1) |  |
| non-Hispanic Asian | 13,193,149 (5.4) | 11,247,636 (5.5) | 1,945,513 (4.8) |  |
| non-Hispanic Other | 12,506,372 (5.1) | 10,494,978 (5.2) | 2,011,393 (4.9) |  |
| Missing | 8,188,790 (3.4) | 7,202,765 (3.5) | 986,025 (2.4) |  |
| **Educational attainment, No. (%)** |  |  |  | **0.042** |
| Less than high school | 14,724,515 (6.0) | 12,385,218 (6.1) | 2,339,297 (5.7) |  |
| High school graduate | 51,599,913 (21.1) | 45,187,651 (22.3) | 6,412,262 (15.7) |  |
| Some college | 93,452,016 (38.3) | 76,782,409 (37.8) | 16,669,607 (40.7) |  |
| College graduate or more | 82,203,703 (33.7) | 67,104,185 (33.0) | 15,099,517 (36.9) |  |
| Missing | 2,019,773 (0.8) | 1,610,977 (0.8) | 408,796 (1.0) |  |
| **Annual household income, No. (%)** |  |  |  | **0.082** |
| <\$50,000 | 94,542,770 (38.7) | 78,963,284 (38.9) | 15,579,486 (38.1) | |
| \$50,000 to \$99,999 | 67,706,737 (27.7) | 55,671,349 (27.4) | 12,035,388 (29.4) | |
| \$100,000+ | 81,239,534 (33.3) | 68,219,406 (33.6) | 13,020,128 (31.8) | |
| Missing | 510,879 (0.2) | 216,402 (0.1) | 294,477 (0.7) |  |
| **Health insurance coverage, No. (%)** |  |  |  | **0.90** |
| No | 20,313,302 (8.3) | 16,824,007 (8.3) | 3,489,295 (8.5) |  |
| Yes | 222,840,218 (91.3) | 185,560,693 (91.4) | 37,279,525 (91.1) |  |
| Missing | 846,399 (0.3) | 685,739 (0.3) | 160,660 (0.4) |  |
| **Urbanicity, No. (%)** |  |  |  | **0.80** |
| Urban | 227,571,581 (93.3) | 189,547,414 (93.3) | 38,024,168 (92.9) |  |
| Rural | 16,428,338 (6.7) | 13,523,026 (6.7) | 2,905,312 (7.1) |  |
| **Census region, No. (%)** |  |  |  | **0.90** |
| Northeast | 42,198,193 (17.3) | 34,688,990 (17.1) | 7,509,202 (18.3) |  |
| Midwest | 51,466,171 (21.1) | 42,502,386 (20.9) | 8,963,785 (21.9) |  |
| South | 94,033,712 (38.5) | 78,460,885 (38.6) | 15,572,827 (38.0) |  |
| West | 56,301,844 (23.1) | 47,418,178 (23.4) | 8,883,666 (21.7) |  |
| P-value by Pearson's Chi-squared test with Rao & Scott adjustment or Design-based Kruskal-Wallis test. |  |  |  |  |
| Bold font indicates $p < 0.05$. | | | | |
| The non-Hispanic Other subgroup included American Indian/Alaska Native, Native Hawaiian/Pacific Islander, and multiracial respondents. |  |  |  |  |

**Table 1 (continued).** Weighted summary statistics, overall and by awareness of multi-cancer early detection tests in the United States, 2024 Health Information National Trends Survey.
| Characteristic | Aware of Multi-Cancer Early Detection tests |  |  | p-value |
| --- | --- | --- | --- | --- |
|  | Overall<br>N = 243,999,920 | No<br>N = 203,070,440 (83.2%) | Yes<br>N = 40,929,480 (16.8%) |  |
| **Personal cancer history, No. (%)** |  |  |  | 0.30 |
| No | 219,288,353 (89.9) | 183,236,532 (90.2) | 36,051,821 (88.1) |  |
| Yes | 23,709,806 (9.7) | 19,025,377 (9.4) | 4,684,429 (11.4) |  |
| Missing | 1,001,760 (0.4) | 808,531 (0.4) | 193,229 (0.5) |  |
| **Family cancer history, No. (%)** |  |  |  | 0.40 |
| No | 53,286,509 (21.8) | 43,762,192 (21.6) | 9,524,317 (23.3) |  |
| Not sure | 30,951,209 (12.7) | 26,909,628 (13.3) | 4,041,581 (9.9) |  |
| Yes | 154,386,416 (63.3) | 127,966,197 (63.0) | 26,420,219 (64.6) |  |
| Missing | 5,375,786 (2.2) | 4,432,423 (2.2) | 943,363 (2.3) |  |
| **Number of chronic health conditions, No. (%)** |  |  |  | 0.40 |
| 0 | 99,173,216 (40.6) | 82,071,210 (40.4) | 17,102,006 (41.8) |  |
| 1 | 73,785,690 (30.2) | 62,634,863 (30.8) | 11,150,827 (27.2) |  |
| 2+ | 71,041,013 (29.1) | 58,364,366 (28.7) | 12,676,647 (31.0) |  |
| **Past 12-month patient portal use, No. (%)** |  |  |  | 0.082 |
| 0 | 73,715,660 (30.2) | 62,934,219 (31.0) | 10,781,441 (26.3) |  |
| 1 to 5 times | 103,457,552 (42.4) | 83,987,272 (41.4) | 19,470,280 (47.6) |  |
| 6 or more times | 66,394,591 (27.2) | 55,775,574 (27.5) | 10,619,016 (25.9) |  |
| Missing | 432,117 (0.2) | 373,375 (0.2) | 58,743 (0.1) |  |
| **Health-related social media use, Mean (SD)** | 10.5 (3.6) | 10.4 (3.5) | 11.0 (3.9) | **<0.001** |
| P-value by Pearson's Chi-squared test with Rao & Scott adjustment or Design-based Kruskal-Wallis test. |  |  |  |  |
| Bold font indicates $p < 0.05$. | | | | |
| The non-Hispanic Other subgroup included American Indian/Alaska Native, Native Hawaiian/Pacific Islander, and multiracial respondents. |  |  |  |  |

Adults who perceived MCED testing as very valuable differed from those in the other category group (not at all valuable/a little valuable/somewhat valuable) across multiple characteristics (Table 2). Adults who perceived MCED testing as very valuable were more likely to be aged 50+ (50.4% vs. 45.9%, *p*<0.001), female (51.9% vs. 45.3%, *p*=0.016), and from minoritized racial/ethnic groups (non-Hispanic Black: 12.4% vs. 9.4%; Hispanic: 19.9% vs. 14.8%, *p*=0.005) compared to those in the other category group. They were also more likely to have family cancer history (66.9% vs. 60.6%, *p*=0.012) and use their patient portal ≥6 times in the past 12 months (31.9% vs. 23.8%, *p*<0.001).

**Table 2.** Weighted summary statistics by perceived personal value of multi-cancer early detection tests in the United States, 2024 Health Information National Trends Survey.

| Characteristic | Perceived Multi-Cancer Early Detection testing as very valuable |  | p-value |
| --- | --- | --- | --- |
|  | No<br>N = 141,250,363 (57.9%) | Yes<br>N = 102,749,557 (42.1%) |  |
| **Age, No. (%)** |  |  | **<0.001** |
| 18-34 | 41,466,278 (29.4) | 19,521,278 (19.0) |  |
| 35-49 | 33,242,538 (23.5) | 29,955,895 (29.2) |  |
| 50-64 | 32,229,738 (22.8) | 31,337,020 (30.5) |  |
| 65-74 | 18,545,321 (13.1) | 12,938,103 (12.6) |  |
| 75+ | 14,089,997 (10.0) | 7,477,417 (7.3) |  |
| Missing | 1,676,489 (1.2) | 1,519,843 (1.5) |  |
| **Sex assigned at birth, No. (%)** |  |  | **0.016** |
| Male | 74,965,742 (53.1) | 47,987,492 (46.7) |  |
| Female | 64,018,643 (45.3) | 53,307,680 (51.9) |  |
| Missing | 2,265,978 (1.6) | 1,454,384 (1.4) |  |
| **Race/ethnicity, No. (%)** |  |  | **0.005** |
| non-Hispanic White | 86,630,269 (61.3) | 56,235,032 (54.7) |  |
| non-Hispanic Black | 13,267,908 (9.4) | 12,693,662 (12.4) |  |
| Hispanic | 20,867,323 (14.8) | 20,417,416 (19.9) |  |
| non-Hispanic Asian | 7,754,357 (5.5) | 5,438,792 (5.3) |  |
| non-Hispanic Other | 7,789,799 (5.5) | 4,716,573 (4.6) |  |
| Missing | 4,940,708 (3.5) | 3,248,082 (3.2) |  |
| **Educational attainment, No. (%)** |  |  | 0.70 |
| Less than high school | 9,078,746 (6.4) | 5,645,769 (5.5) |  |
| High school graduate | 29,857,799 (21.1) | 21,742,114 (21.2) |  |
| Some college | 54,783,789 (38.8) | 38,668,227 (37.6) |  |
| College graduate or more | 46,232,468 (32.7) | 35,971,234 (35.0) |  |
| Missing | 1,297,561 (0.9) | 722,212 (0.7) |  |
| **Annual household income, No. (%)** |  |  | 0.30 |
| <\$50,000 | 57,144,868 (40.5) | 37,397,902 (36.4) | |
| \$50,000 to \$99,999 | 38,123,713 (27.0) | 29,583,024 (28.8) | |
| \$100,000+ | 45,701,360 (32.4) | 35,538,174 (34.6) | |
| Missing | 280,422 (0.2) | 230,457 (0.2) |  |
| **Health insurance coverage, No. (%)** |  |  | 0.40 |
| No | 12,357,025 (8.7) | 7,956,277 (7.7) |  |
| Yes | 128,502,804 (91.0) | 94,337,414 (91.8) |  |
| Missing | 390,533 (0.3) | 455,865 (0.4) |  |
| **Urbanicity, No. (%)** |  |  | 0.20 |
| Urban | 130,964,787 (92.7) | 96,606,794 (94.0) |  |
| Rural | 10,285,576 (7.3) | 6,142,762 (6.0) |  |
| **Census region, No. (%)** |  |  | 0.40 |
| Northeast | 24,791,324 (17.6) | 17,406,869 (16.9) |  |
| Midwest | 31,501,252 (22.3) | 19,964,919 (19.4) |  |
| South | 53,513,787 (37.9) | 40,519,925 (39.4) |  |
| West | 31,444,000 (22.3) | 24,857,844 (24.2) |  |
| P-value by Pearson's Chi-squared test with Rao & Scott adjustment or Design-based Kruskal-Wallis test. |  |  |  |
| Bold font indicates $p < 0.05$. | | | |
| The non-Hispanic Other subgroup included American Indian/Alaska Native, Native Hawaiian/Pacific Islander, and multiracial respondents. |  |  |  |

**Table 2 (continued).** Weighted summary statistics by perceived personal value of multi-cancer early detection tests in the United States, 2024 Health Information National Trends Survey.
| Characteristic | Perceived Multi-Cancer Early Detection testing as very valuable |  | p-value |
| --- | --- | --- | --- |
|  | No<br>N = 141,250,363<br>(57.9%) | Yes<br>N = 102,749,557<br>(42.1%) |  |
| **Personal cancer history, No. (%)** |  |  | 0.40 |
| No | 127,537,695 (90.3) | 91,750,658 (89.3) |  |
| Yes | 13,261,883 (9.4) | 10,447,923 (10.2) |  |
| Missing | 450,785 (0.3) | 550,975 (0.5) |  |
| **Family cancer history, No. (%)** |  |  | **0.012** |
| No | 33,357,144 (23.6) | 19,929,364 (19.4) |  |
| Not sure | 19,400,781 (13.7) | 11,550,427 (11.2) |  |
| Yes | 85,653,110 (60.6) | 68,733,306 (66.9) |  |
| Missing | 2,839,327 (2.0) | 2,536,459 (2.5) |  |
| **Number of chronic health conditions, No. (%)** |  |  | 0.13 |
| 0 | 60,014,531 (42.5) | 39,158,686 (38.1) |  |
| 1 | 41,194,130 (29.2) | 32,591,560 (31.7) |  |
| 2+ | 40,041,702 (28.3) | 30,999,311 (30.2) |  |
| **Past 12-month patient portal use, No. (%)** |  |  | **<0.001** |
| 0 | 45,940,703 (32.5) | 27,774,956 (27.0) |  |
| 1 to 5 times | 61,431,142 (43.5) | 42,026,411 (40.9) |  |
| 6 or more times | 33,586,295 (23.8) | 32,808,296 (31.9) |  |
| Missing | 292,223 (0.2) | 139,894 (0.1) |  |
| **Health-related social media use, Mean (SD)** | 10.5 (3.7) | 10.5 (3.4) | 0.13 |
| P-value by Pearson's Chi-squared test with Rao & Scott adjustment or Design-based Kruskal-Wallis test. |  |  |  |
| Bold font indicates $p < 0.05$. | | | |
| The non-Hispanic Other subgroup included American Indian/Alaska Native, Native Hawaiian/Pacific Islander, and multiracial respondents. |  |  |  |

### Multivariable Analysis of MCED Test Awareness

In adjusted analyses (Table 3), Hispanic ethnicity and health-related social media use were significantly associated with MCED test awareness. Hispanic adults had 29% higher prevalence of awareness compared to non-Hispanic White adults (aPR = 1.29, 95% CI: 1.01-1.65). Additionally, each unit increase in health-related social media use was associated with 4% higher prevalence of awareness (aPR = 1.04, 95% CI: 1.01-1.06). The other demographic, socioeconomic, and health-related factors showed no significant adjusted associations with MCED awareness.

**Table 3.** Factors associated with awareness and perceived personal value of multi-cancer early detection tests in the United States, 2024 Health Information National Trends Survey.

|  | Awareness of Multi-Cancer<br>Early Detection tests |  | Perceived Multi-Cancer Early<br>Detection testing as very valuable |  |
| --- | --- | --- | --- | --- |
| *Characteristic* | *Adjusted PR (95% CI)* | *p value* | *Adjusted PR (95% CI)* | *p value* |
| **Age** |  |  |  |  |
| 18-34 (reference) |  |  |  |  |
| 35-49 | 0.86 (0.64, 1.16) | 0.32 | **1.43 (1.20, 1.70)** | **<0.001** |
| 50-64 | 1.11 (0.84, 1.46) | 0.48 | **1.48 (1.25, 1.76)** | **<0.001** |
| 65-74 | 1.07 (0.77, 1.47) | 0.70 | **1.26 (1.04, 1.52)** | **0.019** |
| 75+ | 1.08 (0.75, 1.57) | 0.66 | 1.11 (0.87, 1.40) | 0.40 |
| **Sex assigned at birth** |  |  |  |  |
| Male (reference) |  |  |  |  |
| Female | 0.88 (0.73, 1.07) | 0.22 | 1.10 (1.00, 1.22) | 0.057 |
| **Race/ethnicity** |  |  |  |  |
| non-Hispanic White (reference) |  |  |  |  |
| non-Hispanic Black | 1.09 (0.83, 1.43) | 0.53 | **1.31 (1.15, 1.50)** | **<0.001** |
| Hispanic | **1.29 (1.01, 1.65)** | **0.039** | **1.40 (1.22, 1.60)** | **<0.001** |
| non-Hispanic Asian | 0.90 (0.51, 1.57) | 0.70 | 1.18 (0.92, 1.52) | 0.20 |
| non-Hispanic Other | 1.03 (0.67, 1.57) | 0.90 | 1.04 (0.80, 1.35) | 0.79 |
| **Educational attainment** |  |  |  |  |
| Less than high school (reference) |  |  |  |  |
| High school graduate | 0.83 (0.54, 1.26) | 0.38 | 1.12 (0.91, 1.39) | 0.29 |
| Some college | 1.16 (0.78, 1.71) | 0.46 | 1.06 (0.86, 1.30) | 0.60 |
| College graduate or more | 1.28 (0.86, 1.90) | 0.22 | 1.10 (0.90, 1.36) | 0.35 |
| **Annual household income** |  |  |  |  |
| <\$50,000 (reference) | | | | |
| \$50,000 to \$99,999 | 1.02 (0.80, 1.29) | 0.90 | 1.08 (0.95, 1.23) | 0.26 |
| \$100,000+ | 0.88 (0.69, 1.13) | 0.32 | 1.04 (0.91, 1.20) | 0.53 |
| **Health insurance coverage** |  |  |  |  |
| No (reference) |  |  |  |  |
| Yes | 0.97 (0.70, 1.35) | 0.87 | 1.01 (0.84, 1.23) | 0.88 |
| **Urbanicity** |  |  |  |  |
| Urban (reference) |  |  |  |  |
| Rural | 1.15 (0.79, 1.67) | 0.47 | 0.95 (0.77, 1.17) | 0.63 |
| **Census region** |  |  |  |  |
| Northeast (reference) |  |  |  |  |
| Midwest | 0.98 (0.72, 1.34) | 0.89 | 0.96 (0.81, 1.14) | 0.62 |
| South | 0.91 (0.69, 1.21) | 0.53 | 1.03 (0.89, 1.19) | 0.70 |
| West | 0.87 (0.63, 1.19) | 0.38 | 1.07 (0.91, 1.26) | 0.41 |
| **Personal cancer history** |  |  |  |  |
| No (reference) |  |  |  |  |
| Yes | 1.15 (0.92, 1.46) | 0.23 | 1.06 (0.93, 1.20) | 0.37 |
| **Family cancer history** |  |  |  |  |
| No (reference) |  |  |  |  |
| Not sure | 0.75 (0.50, 1.13) | 0.17 | 1.04 (0.85, 1.26) | 0.70 |
| Yes | 0.99 (0.79, 1.24) | 0.94 | **1.17 (1.03, 1.34)** | **0.014** |
| **Number of chronic health conditions** |  |  |  |  |
| 0 (reference) |  |  |  |  |
| 1 | 0.86 (0.68, 1.08) | 0.19 | 1.10 (0.97, 1.23) | 0.14 |
| 2 + | 1.01 (0.81, 1.26) | 0.93 | 1.07 (0.94, 1.22) | 0.30 |
| **Past 12-month patient portal use** |  |  |  |  |
| None (reference) |  |  |  |  |
| 1 to 5 times | 1.22 (0.96, 1.55) | 0.10 | 1.02 (0.90, 1.16) | 0.75 |
| 6 or more times | 1.04 (0.78, 1.38) | 0.80 | **1.23 (1.06, 1.42)** | **0.006** |
| **Health-related social media use** | **1.04 (1.01, 1.06)** | **0.002** | 1.00 (0.99, 1.01) | 0.97 |
| PR: prevalence ratio; CI: confidence interval. |  |  |  |  |
| The non-Hispanic Other subgroup included American Indian/Alaska Native, Native Hawaiian/Pacific Islander, and multiracial respondents. |  |  |  |  |
| Results were pooled across ten imputed datasets. |  |  |  |  |
| Bold font indicates $p < 0.05$. | | | | |

### Multivariable Analysis of Perceived MCED Value

Multiple factors were significantly associated with perceiving MCED tests as very valuable in adjusted analyses (Table 3). Adults aged 35-49 (aPR = 1.43, 95% CI: 1.20-1.70), 50-64 (aPR = 1.48, 95% CI: 1.25-1.76), and 65-74 years (aPR = 1.26, 95% CI: 1.04-1.52) were significantly more likely to perceive MCED tests as very valuable compared to those aged 18-34 years. Non-Hispanic Black (aPR = 1.31, 95% CI: 1.15-1.50) and Hispanic (aPR = 1.40, 95% CI: 1.22-1.60) adults were significantly more likely to perceive MCED tests as very valuable compared to non-Hispanic White adults. Health-related factors were also significantly associated with perceived value. Adults with family cancer history were 17% (aPR = 1.17, 95% CI: 1.03-1.34) more likely to perceive MCED tests as very valuable compared to those without family cancer history. Frequent patient portal users (≥6 times in the past year) were 23% (aPR = 1.23, 95% CI: 1.06-1.42) more likely to perceive MCED tests as very valuable compared to non-users.

## Discussion

MCED testing has the potential to reduce the U.S. cancer burden by detecting multiple cancer signals early (2,3). However, at $949 per test without insurance coverage, understanding population awareness and perceived value is crucial for informing policy decisions and clinical implementation (2–6). Only 17% of U.S. adults were aware of MCED tests, yet 42% perceived having an MCED test as very valuable. This indicates a substantial awareness-value gap. Hispanic individuals and those with high health-related social media use showed greater MCED test awareness compared to their White, lower health-related social media use counterparts. Perceived value was highest among older adults, minoritized racial/ethnic groups, individuals with family cancer history, and frequent patient portal users. These patterns suggest that while awareness remains limited and concentrated in specific populations, perceived value is high among those who may benefit most from early cancer detection (6,37,38).

Our hypothesis that MCED test awareness would be below 20% was confirmed, which is plausible given that MCED tests lack FDA approval and are not covered by health insurance plans (2,3). However, our hypothesis about demographic predictors of awareness was largely unsupported. In adjusted analyses, awareness did not significantly differ by age, socioeconomic status, personal or family cancer history, or urbanicity. These limited associations contrast with patterns exhibited for a similar technology—genetic testing—where education, income, and personal cancer history are strong predictors (12–14,16,17,19,20). However, our findings align with recent MCED interest studies (7,8). A large urban health system study found that age, education, and health insurance coverage did not predict MCED testing interest (8). Similarly, a national study of adults aged 45-80 reported no associations between MCED willingness and age, household income, metropolitan status, or personal cancer history (7).

We found that Hispanic ethnicity and health-related social media use were associated with higher MCED test awareness. While literature specific to MCED test awareness among Hispanic/Latino populations is limited, our finding that Hispanic individuals had higher awareness than non-Hispanic White individuals warrants further investigation. Although the HINTS 7 dataset did not include measures of generational status, immigration history, or acculturation levels, these factors may be critical to understanding awareness patterns within Hispanic/Latino communities (39,40). Research on genetic testing has documented important differences between U.S.-born and non-U.S. born Hispanic/Latino populations, with the Hispanic Community Health Study/Study of Latinos (2018–2020) showing that non-U.S. born individuals perceived genetic testing as more useful than their U.S.-born counterparts (41). Additionally, acculturation levels, language preferences, and generational differences in health information-seeking behaviors could influence how Hispanic/Latino individuals learn about and perceive novel screening technologies like MCED tests (39,40). Future research should collect data on these additional measures to better understand potential heterogeneity within Hispanic/Latino populations and identify which subgroups may have higher MCED test awareness. Such granular data would enable researchers to develop more targeted and culturally appropriate educational strategies.

Our finding that health-related social media use was associated with greater MCED test awareness aligns with established literature linking internet use to health technology awareness (12,13,19). Given this association, ensuring equitable access to information about this emerging cancer screening technology will require better understanding of how MCED tests are being discussed on social media platforms (42). Additionally, we found that greater patient portal use was significantly associated with higher perceived value of MCED tests. This association may reflect that individuals who frequently use patient portals generally place greater value on novel health technologies (21). This finding raises important questions for future studies about how to reach patients with less patient portal engagement.

### Strengths, Limitations, and Future Research Directions

This study has several notable strengths. First, by incorporating HINTS 7’s complex survey design and analytic weights, our findings generalize to the noninstitutionalized U.S. adult population. Second, we used a composite measure of health-related social media use that captured multiple dimensions of online health engagement (29) and had acceptable reliability (α = 0.71) (43). Third, given that the prevalences of MCED test awareness (16.8%) and perceived value (42.1%) were greater than 10% (44), we estimated prevalence ratios rather than odds ratios. This methodological choice prevented the overestimation of effect sizes that occurs with logistic regression (44). Thus, we provide more accurate and interpretable association measures for health communication scientists, genetic counselors, and clinicians.

Despite its strengths, this study has several limitations. First, the analysis was restricted to the U.S. population, limiting generalizability to countries with different healthcare systems and demographic compositions. Second, this study relied on self-reported measures, which may be subject to recall, misclassification, and social desirability biases. Third, unmeasured confounders may influence the observed associations. For example, we analyzed race/ethnicity as a demographic factor, but this measure is a social construct that serves as a proxy for structural mechanisms such as neighborhood disadvantage (45–47). Future studies should test whether the structural factors impact MCED test awareness and perceived value. Additionally, risk factors such as Ashkenazi ancestry were not available in HINTS 7 but may influence MCED awareness and perceived value. Finally, the observed associations should not be interpreted as causal given the study’s cross-sectional design. Repeated cross-sectional analyses would be valuable for monitoring temporal trends in MCED awareness as these tests become more widely available. Longitudinal studies are also needed to understand how MCED awareness translates to actual MCED test uptake over time.

## Conclusion

Using HINTS 7 data, we provide nationally representative estimates of MCED test awareness and perceived value among U.S. adults. These findings highlight potential opportunities and challenges for the equitable implementation of this emerging cancer screening technology. While only 17% of U.S. adults were aware of MCED tests, 42% perceived them as very valuable, indicating substantial potential demand if awareness barriers are addressed. The higher awareness among Hispanic individuals suggests the need for evolving pathways for the dissemination of health technology information, which warrants further investigation. In conclusion, as MCED tests move toward FDA approval and potential insurance coverage, this descriptive report can inform data-driven strategies to ensure equitable access.

## Data Availability

The data supporting these findings are publicly available on the Health Information National Trends Survey website.

https://hints.cancer.gov/data/download-data.aspx

## Author contributions

**Jemar R. Bather:** Conceptualization, Methodology, Software, Formal analysis, Investigation, Data Curation, Writing (Original Draft), Project administration. **Melody S. Goodman:** Methodology, Validation, Investigation, Resources, Writing (Review & Editing). **Daniel Chavez-Yenter:** Validation, Investigation, Writing (Review & Editing). **José A. Pagán:** Validation, Investigation, Writing (Review & Editing). **Kimberly A. Kaphingst:** Conceptualization, Investigation, Data Curation, Writing (Review & Editing).

## Notes

### Competing Interest Statement

The authors have declared no competing interest.

### Funding Statement

This research did not receive any specific grant from funding agencies in the public, commercial, or not-for-profit sectors.

